# Everolimus improves the efficacy of dasatinib in PDGFRα-driven glioma

**DOI:** 10.1101/2019.12.28.19015974

**Authors:** Zachary Miklja, Viveka Nand Yadav, Rodrigo T. Cartaxo, Ruby Siada, Chase C. Thomas, Jessica R. Cummings, Brendan Mullan, Stefanie Stallard, Alyssa Paul, Amy K. Bruzek, Kyle Wierzbicki, Tao Yang, Taylor Garcia, Ian Wolfe, Marcia Leonard, Patricia L. Robertson, Hugh J.L. Garton, Daniel R. Wahl, Hemant Parmar, Jann N. Sarkaria, Cassie Kline, Sabine Mueller, Theodore Nicolaides, Chana Glasser, Sarah E.S. Leary, Sriram Venneti, Chandan Kumar-Sinha, Arul M. Chinnaiyan, Rajen Mody, Manjunath P. Pai, Timothy N. Phoenix, Bernard L. Marini, Carl Koschmann

## Abstract

**Background:** Pediatric and adult high-grade glioma (HGG) frequently harbor *PDGFRA* alterations. We hypothesized that co-treatment with everolimus may improve the efficacy of dasatinib in PDGFRα-driven glioma through combinatorial synergism and increased tumor accumulation of dasatinib.

**Methods:** Dose response, synergism studies, P-gp inhibition and pharmacokinetic studies were performed on in vitro and in vivo human and mouse models of HGG. Six patients with recurrent PDGFRα-driven glioma were treated with dasatinib and everolimus.

**Results:** Dasatinib effectively inhibited the proliferation of mouse and human primary HGG cells with a variety of *PDGFRA* alterations. Dasatinib exhibited synergy with everolimus in the treatment of HGG cells at low nanomolar concentrations of both agents, with reduction in mTOR signaling that persists after dasatinib treatment alone. Prolonged exposure to everolimus significantly improved the CNS retention of dasatinib and extended survival of PPK tumor bearing mice. Pediatric patients (n=6) with glioma tolerated this combination without significant adverse events. Recurrent patients (n=4) demonstrated median overall survival of 8.5 months.

**Conclusion:** Efficacy of dasatinib treatment of PDGFRα-driven HGG is improved with everolimus and suggests a promising route for improving targeted therapy for this patient population.

**Trial Registration:** ClinicalTrials.gov NCT03352427

**Funding:** The authors thank the patients and their families for participation in this study. CK is supported by NIH/NINDS K08-NS099427-01, the University of Michigan Chad Carr Pediatric Brain Tumor Center, the Chad Tough Foundation, Hyundai Hope on Wheels, Catching up With Jack, Prayers from Maria Foundation, U CAN-CER VIVE FOUNDATION, Morgan Behen Golf Classic, and the DIPG Collaborative. The PEDS-MIONCOSEQ study was supported by grant 1UM1HG006508 from the National Institutes of Health Clinical Sequencing Exploratory Research Award (PI: Arul Chinnaiyan).

## INTRODUCTION

High-grade gliomas (HGGs) are common and aggressive pediatric and adult brain tumors. Median overall survival (OS) of adult patients diagnosed with glioblastoma (GBM), grade IV glioma, is 12.6 months ((1)), and median OS for pediatric HGG is 14.1 months (2). Established therapies for adult and pediatric HGG are not currently targeted to the unique molecular attributes of each tumor, and patients frequently consider experimental therapies after up-front radiation and at recurrence.

One of the most frequently altered genes in HGG is platelet-derived growth factor receptor alpha (*PDGFRA). PDGFRA* is one of two PDGF receptor subunits, and it interacts with at least four PDGF ligands (3). It has been shown that PDGF activation induces multiple cellular activities including cell proliferation, transformation, migration, and survival (3). *PDGFRA* is mutated or amplified in 12% of adult GBM and 21% of pediatric HGG (4, 5). Within childhood HGG, *PDGFRA* mutation is correlated with older age (10-30 years), while amplification is seen more commonly in younger patients (5-10 years) (5). *PDGFRA* alterations are drivers of aggressive glioma behavior and are associated with worse prognoses in pediatric non-brainstem HGG and adult anaplastic astrocytoma (WHO grade III glioma) (5, 6).

Pre-clinically, there are multiple effective agents targeting *PDGFRA* in glioma, including the tyrosine kinase inhibitor (TKI) dasatinib (5, 7). Unfortunately, monotherapy with dasatinib has failed to improve outcomes in adult HGG, even when gliomas were selected for by PDGFRα overexpression by IHC (8). The failure of dasatinib may be related to its use as a single agent. Recent data in other solid tumors has demonstrated synergism between TKIs and the mTOR inhibitor everolimus, including the clinical success of lenvatinib and everolimus for renal cell carcinoma (9). Osteosarcoma pre-clinical models treated with sorafenib overcame resultant upregulation of mTORC1 (mTOR complex 1) through co-treatment with everolimus (10).

Additionally, treatment with dasatinib for CNS tumors may be limited by CNS penetration. Based on its intrinsic qualities (i.e. lipophilicity, size, and protein binding), dasatinib is a promising agent for CNS penetration (11). While the ability of dasatinib to penetrate the blood-brain barrier (BBB) is promising, its retention is limited by CSF efflux proteins P-glycoprotein (P-gp) and breast cancer resistance protein (BCRP) (12). Recent work has demonstrated that co-administration of TKIs with agents that inhibit P-gp and Bcrp1 can improve the brain retention of the TKI (12-14). Everolimus has been shown to improve the CNS retention of the TKI vandetanib (13).

To date, co-administration of dasatinib and everolimus has never been tested in CNS tumor models or administered to human patients with PDGFRα-driven gliomas. In this study, we assessed whether co-treatment of dasatinib with everolimus could improve the efficacy and CNS retention of dasatinib in PDGFRα-driven gliomas in both pre-clinical models and human patients. We found that everolimus acts synergistically with dasatinib in PDGFRα-driven glioma. We also found that sustained everolimus administration results in significant increases in tumor concentrations of dasatinib in a genetically-engineered HGG animal model. Preliminary data from six patients treated establishes promising feasibility. Our findings represent a novel route for enhancing the efficacy of TKI precision therapy for pediatric and adult HGG.

## RESULTS

### Generation of TP53, PDGFRA and H3K27M mutant (PPK) glioma primary cell cultures and in vitro treatment with dasatinib

In order to study the utility of dasatinib in PDGFRα-driven HGG, we adapted an intra-uterine electroporation (IUE) mouse model (15) (Figure 1A). We induced glioma in mice by injecting plasmids encoding: [1] PBase; [2] PB-CAG-DNp53-Ires-Luciferase (“TP53”); [3] PB-CAG-*Pdgfra*D824V-Ires-eGFP (“*PDGFRA* D842V”); and [4] PB-CAG-H3.3 K27M-Ires-eGFP (“H3K27M”) into the lateral ventricles (forebrain) of embryos in CD1 mice at E13.5 days. Transfection efficiency and tumor growth were monitored by in vivo imaging of luminescence (Figure 1A). With the injection of PPK plasmids, mice developed high-grade, invasive glial tumors (Supplemental Figure 1A) with a high penetrance (>90%) and a median survival of 52 days. IHC analysis confirmed up-regulation of total PDGFRα, tumor-specific expression of H3.3K27M, and reduced expression of H3.3K27me3, as expected for H3.3K27M mutant tumors (16) (Supplemental Figure 1B). Multiple TP53, *PDGFRA* and H3K27M (PPK) mutant glioma primary neurosphere cultures were generated from dissociation of intra-cranial tumor tissue from these mice (Figure 1A), with confirmed mutations in cultured neurospheres (Supplemental Figure 1C).

**Figure 1:**
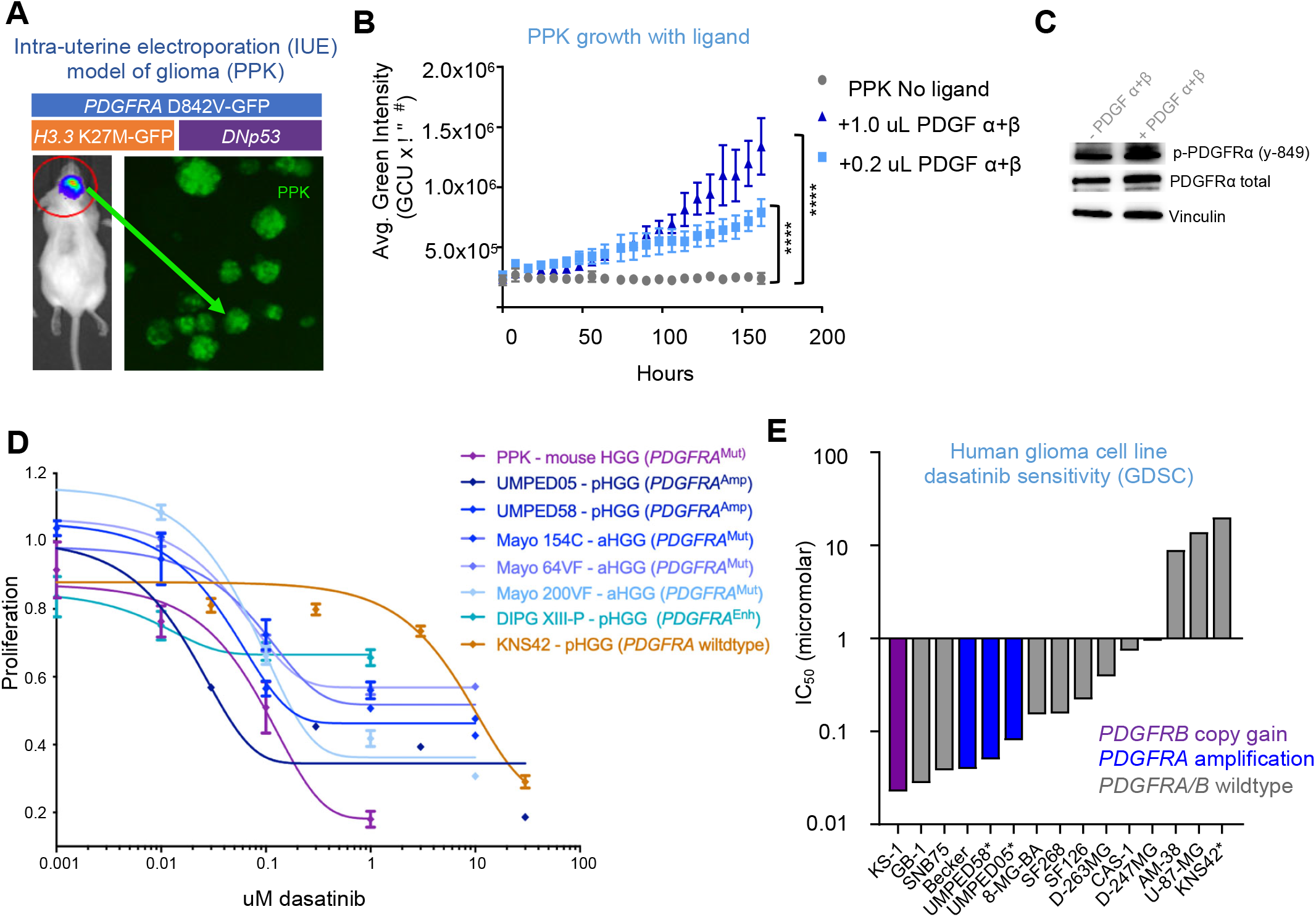
PDGFRα-driven glioma demonstrates sensitivity to dasatinib in vitro **A)** HGG cells are generated from an intra-uterine electroporation (IUE) model with plasmids of *TP53, PDGFRA* and H3K27M mutations. **B)** Proliferation of IUE HGG cells in response to adding PDGF ligand α and β in vitro (**** *P* ≤ 0.0001 by Dunnett’s multiple comparisons test). [n=3 technical replicates]. **C)** Western blot analysis of p-PDGFRα and total PDGFRα in response to PDGF ligand addition. **D)** Dose response curve of PPK neurospheres, cultured cells from six human PDGFRα-driven glioma cell lines, and cells from one *PDGFRA*-wildtype glioma cell line to treatment with dasatinib. [n=3 technical replicates]. **E)** Plot of glioma cell culture sensitivity to dasatinib from Genomics of Drug Sensitivity in Cancer (GDSC). Analysis of *PDGFRA, PDGFA, PDGFRB and PDGFB* DNA-alterations is referenced (*non-GDSC data generated from primary cell culture treatment for this study). [Data represent mean ± SEM for B and D].

Previous studies have demonstrated that tumor cells driven by *PDGFRA* D842V mutation may be relatively PDGF α and β ligand-insensitive (as compared to *PDGFRA* amplification) (7). However, we found a dose-dependent increase in proliferation when cells were supplemented with ligands PDGF α and β (Figure 1B). Cell cultures showed strong expression of activated [p-PDGFRα (y-849)] and total PDGFRα, which was not significantly different with addition of ligand (Figure 1C and Supplemental Figure 1D). UMPED58, brainstem HGG primary cell culture with *PDGFRA* amplification, also demonstrated increased proliferation when grown with ligand PDGF α and β (*P* ≤ 0.0001) (Supplemental Figure 1E).

We tested in vitro dasatinib treatment on mouse PPK and several other human pediatric HGG with *PDGFRA* alteration: two pediatric HGG (pHGG) cell cultures with *PDGFRA* amplification [UMPED58 and UMPED05 (5)], three *PDGFRA*-mutant adult HGG (aHGG) cultures derived from patient-derived xenografts (PDXs) (17), one pHGG cell culture with enhancer-mediated *PDGFRA* up-regulation [DIPG-XIII-P (18)], and one pediatric HGG cell culture without any growth factor receptor alteration [KNS42] as a control (5). PPK cells with *PDGFRA* D842V mutation demonstrated sensitivity to monotherapy with dasatinib with an IC50 of 100 nM (Figure 1D). All the pHGG cell cultures also demonstrated different degrees of nanomolar sensitivity to dasatinib; UMPED05 (*PDGFRA* amplification) showed maximum sensitivity and KNS42 with wildtype *PDGFRA* was the least sensitive (Figure 1D). We analyzed publicly available human glioma cell culture dasatinib treatment data (n=11) from the Genomics of Drug Sensitivity in Cancer (GDSC) (19). Glioma cell cultures demonstrated promising sensitivity to dasatinib in general (Supplemental Figure 2). Dasatinib demonstrated improved efficacy in glioma cell cultures with *PDGFR* alterations (Figure 1E). Collectively, this data demonstrated that dasatinib effectively inhibits proliferation in mouse and human HGG with various *PDGFR* alterations.

**Figure 2:**
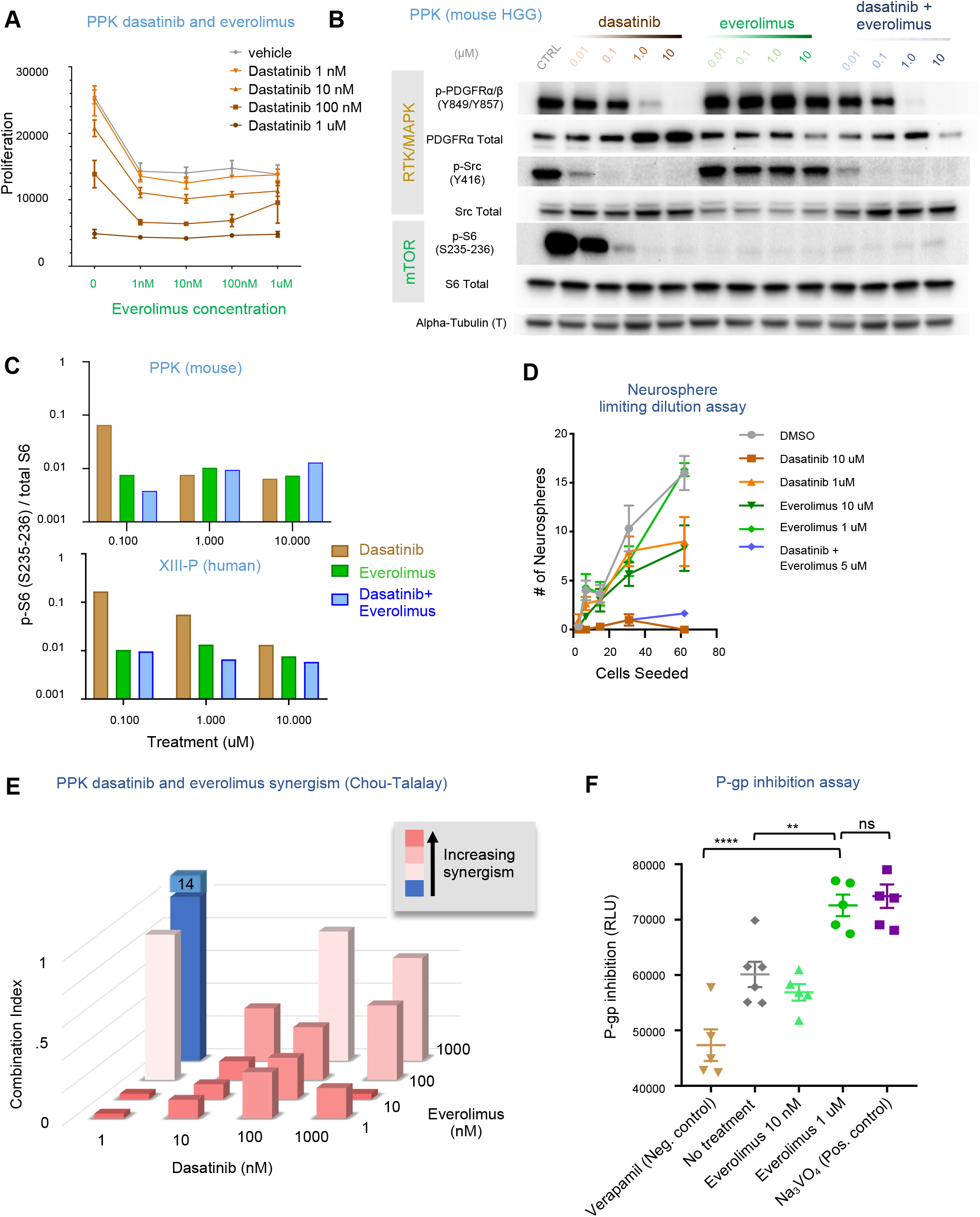
Addition of everolimus provides synergism and strong p-glycoprotein (P-gp) blockade **A)** Viability of IUE HGG neurospheres in response to various combinations of dasatinib and everolimus. [n=3 technical replicates]. **B)** Western blot analysis of p-PDGFRα/β (Y849/Y857), p-Src (Y416), and p-S6 (S235-236) expression in *PDGFRA*-amplified mouse HGG cells treated with dasatinib or everolimus monotherapy, or co-administration of dasatinib and everolimus. In the co-administration condition, drugs were administered at equal doses (doses indicated in text above figure). **C)** Quantification of western blot analysis for mouse HGG cell line (PPK) and a human *PDGFRA*-enhanced cell line (XIII-P), demonstrating greater reduction in p-S6 expression with co-administration of dasatinib and everolimus than with monotherapy of either drug alone at 0.1 uM in the mouse cell line and all administered doses in the human cell line. **D)** Limiting dilution assay performed on IUE PPK neurospheres treated with either 1uM or 10uM doses of dasatinib or everolimus, or dasatinib and everolimus together (5uM). [n=3 technical replicates]. **E)** Everolimus and dasatinib synergism combination index assessment. **F)** Plot of P-gp inhibition using in vitro assay with controls (higher numbers = greater P-gp inhibition), displaying higher P-gp inhibition in the 1uM dose of everolimus condition than in the negative control or no treatment conditions (**** *P* ≤ 0.0001 and ** *P* = 0.0045 by Tukey’s multiple comparisons test, respectively). [Data represent mean ± SEM for A, D, and F].

### Addition of everolimus provides synergism to dasatinib treatment of pHGG cells

Previous work has established that TKIs may have improved efficacy in combination with the mTOR inhibitor everolimus (9, 10). To assess if the addition of everolimus can improve the efficacy of dasatinib via inhibition of the mTOR pathway, western blot analysis was performed on mouse (PPK) and human (DIPG-XIII-P) cells treated with dasatinib and/or everolimus to assess activity of receptor tyrosine kinase (RTK)/MAP-kinase (MAPK)/mTOR pathways (Figure 2B). Treatment with dasatinib alone showed reduced expression of p-PDGFRα/β (Y849/Y857) in a dose dependent manner and RTK/MAPK targets p-Src (Y416), p-ERK1/2 (Thr202/Tyr204), even at the lowest dose (0.01 µM) (Figure 2B and Supplemental Figure 3). Everolimus effectively inhibited the mTOR pathway [p-S6 (S235-236)] at the lowest dose (0.01 µM). The combination of everolimus and dasatinib led to dual inhibition of mTOR (p-S6) and RTK/MAPK (p-SRC) pathways at 0.01 nM dose, (Figure 2B). Quantification of western blot data showed a reduction in residual mTOR (p-S6) pathway after dasatinib treatment alone (Figure 2C and Supplemental Figure 4). A neurosphere limiting dilution assay of IUE PPK cells was performed in the uM range for both dasatinib and everolimus, indicating no enhancement of effect beyond cytotoxicity seen in proliferation assays at this dose; suggesting these agents do not have added impact on stemness of cultured cells (Figure 2D and Supplemental Figure 5A). Using the Chou-Talalay median-effect method for drug combination analysis, everolimus and dasatinib demonstrated synergistic effect at nearly all dose combinations, including single nanomolar doses of both agents (Figure 2E) (20).

**Figure 3:**
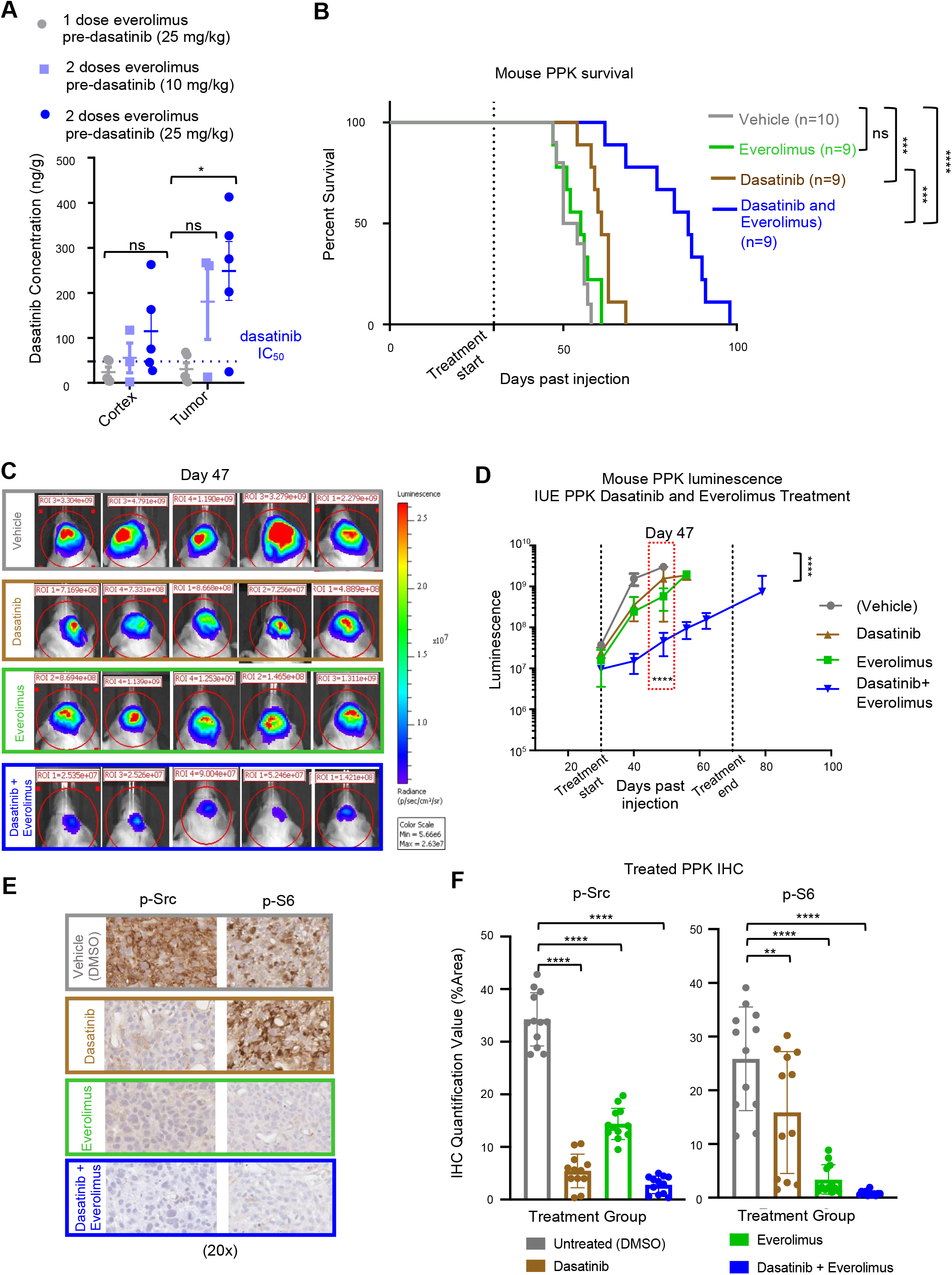
Sustained exposure of everolimus improves dasatinib tumor concentration and efficacy in mouse model **A)** Pharmacokinetic data documenting increase in dasatinib levels found in tumors with additional 24 hour everolimus pre-dasatinib dose of either 5 mg/kg or 10 mg/kg (* *P* ≤ 0.05 by Dunnett’s multiple comparisons test). [n=5 replicates]. **B)** Survival curve data for mice with *PDGFRA-*amplified HGG via IUE demonstrates that median survival for the control condition was 52 days post IUE injection (n=10); the everolimus condition (5 mg/kg, n=9) was 55 days; the dasatinib condition (10 mg/kg, n=9) was 61 days; and the dasatinib and everolimus condition (10 mg/kg and 5 mg/kg, respectively, n=9) was 86 days (*ns P =* 0.3378, *** *P* ≤ 0.0005, **** *P* ≤ 0.0001 by the log-rank test, respectively). **C)** Representative bioluminescence images of PPK mouse tumors (vehicle, n=10; dasatinib, n=9; everolimus, n=9; dasatinib+everolimus, n=10) at 47 days post IUE injection, displaying lower average luminescence in the co-treatment group than in both groups receiving monotherapy of dasatinib or everolimus. **D)** IUE PPK bioluminescence tumor monitor growth data with statistical significance between dasatinib+everolimus and DMSO treatment groups at 47 days post IUE injection (**** *P* ≤ 0.0001 by Tukey’s multiple comparisons test.). **E)** In vivo immunohistochemistry images at 20x magnification of p-Src and p-S6 expression level documenting a reduction with everolimus and dasatinib treatment in comparison to non-treated tumors. [images representative of n=3 biological replicates]. **F)** IHC quantification data for treated PPK mice demonstrates statistical significance of p-Src and p-S6 expression levels between tumors treated with dasatinib and everolimus and non-treated tumors (**** *P* ≤ 0.0001, ** *P* = 0.0065 by Dunnett’s multiple comparisons test). [n=3 animals per treatment group, 4 images per animal]. [Data represent mean ± SEM for A, D, and F].

**Figure 4:**
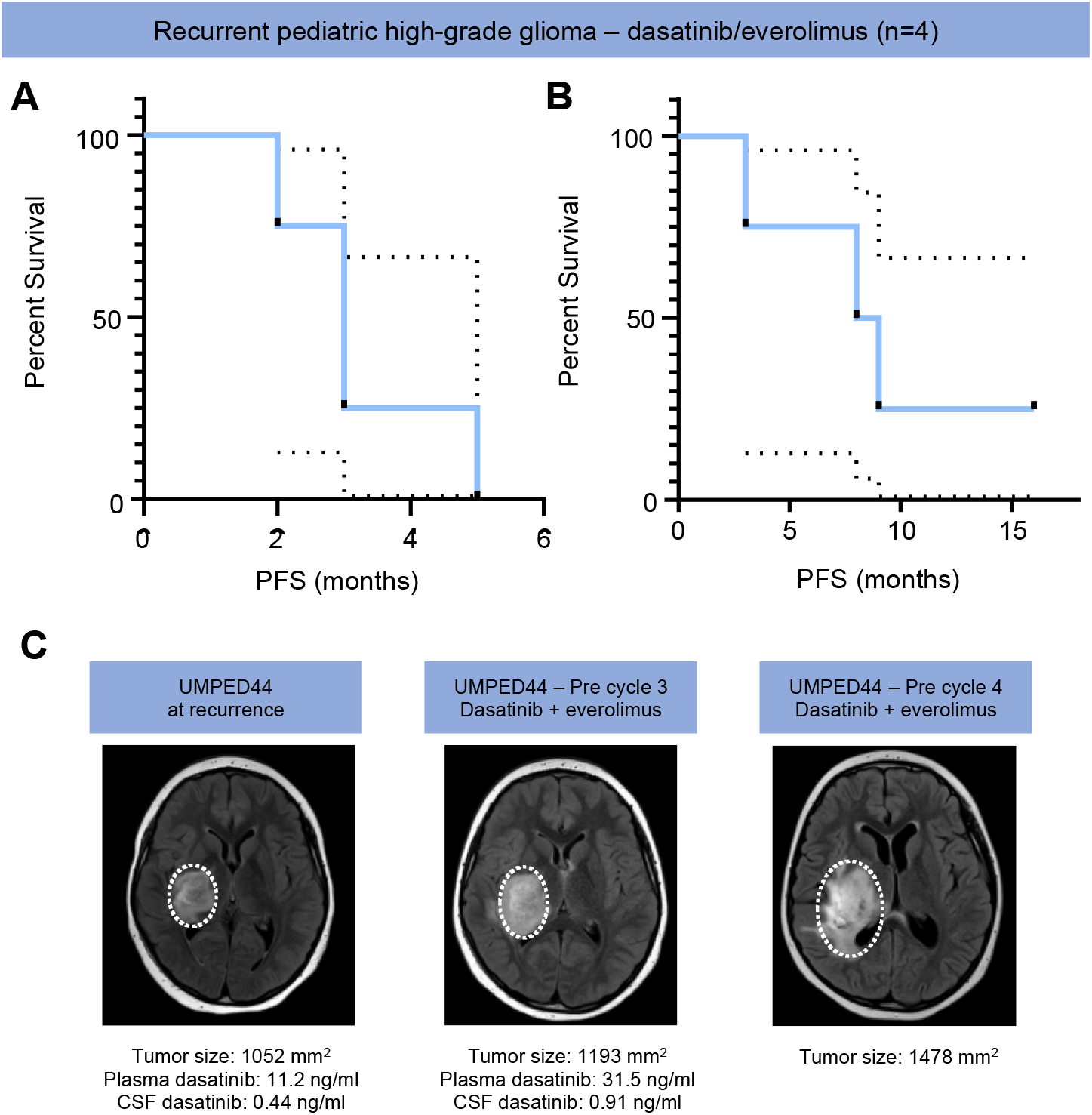
Human survival curves and treatment of UMPED44 with relapsed *PDGFRA*-amplified HGG **A)** Progression-free survival curve for patients with recurrent PDGFRα-driven HGG treated with dasatinib and everolimus (n=4). Dashed lines represent 95% confidence interval. **B)** Overall survival curve for patients with recurrent PDGFRα-driven HGG treated with dasatinib and everolimus (n=4). Dashed lines represent 95% confidence interval. **C)** UMPED44 axial FLAIR MRI at baseline, pre cycle 3, and pre cycle 4 (MR spectroscopy at cycle 4 confirmed tumor progression).

**Figure 5:**
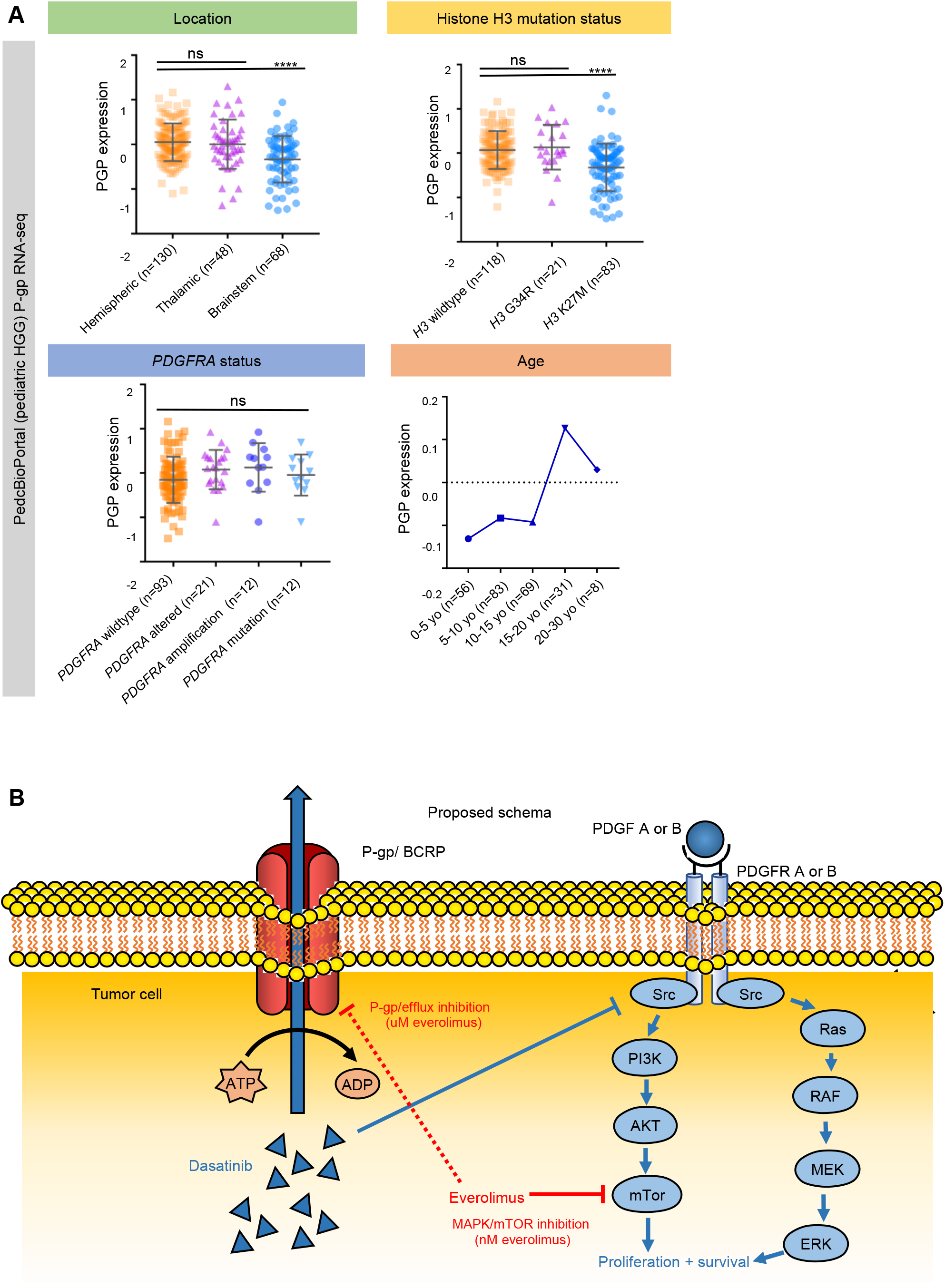
P-gp expression in human pediatric HGG and proposed schema. **A)** PedcBioPortal (pediatric HGG) P-gp RNA-seq data by location (upper left), histone H3 mutation status (upper right), *PDGFRA* status (lower left), and age (lower right) (**** *P* ≤ 0.0001 by Tukey’s multiple comparisons test). Data presented in upper two graphs and lower left graph reflect mean ± SEM. **B)** Proposed schema of dasatinib and everolimus targeting of PDGF-driven HGG. Red line depicting inhibition of P-gp/BCRP by everolimus is dashed to reflect the higher concentration (uM) required to achieve P-gp inhibition than to achieve MAPK/mTOR inhibition (nM).

### Everolimus provides P-gp blockade at higher dose

Previous work has established that dasatinib is a substrate for efflux proteins P-gp and BCRP (21-24). As P-gp is likely the predominant substrate affecting CNS efflux (14), we focused our attention on P-gp. To assess the P-gp inhibitory effects of everolimus, we performed an in vitro P-gp inhibition assay. Everolimus fully blocked P-gp activity at 1 µM at four hours (Figure 2F and Supplemental Figure 3B). Lower doses of everolimus (10 nM) did not produce an effect which was significantly different from non-treated cells (Figure 2F). The effect at 1 µM concentrations was similar to slightly lower than previous levels of everolimus (3-5 µM) necessary for in vitro P-gp inhibition in Madin-Darby canine kidney (MDCK) cells over-expressing multi-drug resistance genes (P-gp and BCRP) (25).

### Pharmacokinetic (PK) analysis of dasatinib and everolimus CNS retention

We then proceeded to test the effect of everolimus on CNS concentrations of dasatinib in non-tumor bearing mice. Mice were treated with dasatinib (10 mg/kg tail vein injection) with or without a single dose of everolimus (5 mg/kg oral gavage administration) two hours prior. Mice were sacrificed at one, two, four, and seven-hour time points (n=3 each) for collection of serum, brainstem and cortex (Supplemental Figure 6A). We did separate analysis of brain and brainstem PK because previous literature and clinical experience suggest that the blood-brain barrier (BBB) is more restrictive in the brainstem (26). No statistically significant differences were found for dasatinib concentrations between the two conditions (dasatinib alone and dasatinib with everolimus) within the plasma, cortex, and brainstem (Supplemental Figure 6B-F). Average plasma concentrations of dasatinib were higher in co-treated mice, although this was not statistically significant.

Pharmacokinetic analysis of everolimus monotherapy in non-tumor bearing mice demonstrated an increase in everolimus concentrations (ng/g) within mouse brain tissue over time (Supplemental Figure 7A-B).

### Sustained exposure of everolimus improves dasatinib tumor delivery and increases survival of mice bearing PPK tumor

We then proceeded to our IUE murine model of pHGG to assess whether intra-tumor concentration of dasatinib would be impacted by co-treatment with everolimus. We performed treatment studies in mice with large cortical PPK tumors that met enrollment luminescence criteria (average 32 dpi). As seen in our non-tumor bearing mice, single dose treatment of everolimus did not influence the tumor concentration of dasatinib (Figure 3A). However, with the addition of a second dose of everolimus 24 hours prior to dasatinib, tumor concentrations of dasatinib were increased, in proportion to dasatinib dose (Figure 3A). In additional tumor bearing mice treated with two doses of everolimus prior to dasatinib treatment immediately before sacrificing demonstrated reduced expression of P-gp by IHC (Supplemental Figure 8A-B).

To assess the effect of sustained co-treatment on survival, IUE PPK mice were treated in four separate conditions: vehicle, daily treatment with dasatinib alone (10 mg/kg), daily treatment with everolimus alone (5 mg/kg), and daily co-administration with dasatinib and everolimus (10 mg/kg and 5 mg/kg respectively). Co-treatment with dasatinib and everolimus significantly improved median survival (86 days) compared to dasatinib only (61 days), everolimus only (55 days) and vehicle alone (52 days) (Figure 3B). At 47 days post IUE injection, the dasatinib and everolimus treated PPK mice demonstrated reduced luminescent tumor signals when compared to control mice (Figure 3C-D). We further analyzed tumors from each treatment group for p-Src and p-S6 expression. IHC analysis and quantification showed reductions in p-Src and p-S6 expression following dasatinib and everolimus monotherapy, with retained p-S6 expression in tumors treated with dasatinib alone (Figure 3E-F, and Supplemental Figure 9A-B).

### Treatment of children with relapsed PDGFRα-driven glioma with dasatinib and everolimus

Patients with recurrent HGG have no proven therapies and an expected overall survival of four to six months (27, 28). Based on the promising pre-clinical data with dasatinib and everolimus, we moved forward with pursuit of clinical treatment with daily dasatinib and everolimus for PDGFRα-driven glioma in human patients. We treated a series of children and young adults with confirmed PDGFRα-driven glioma with continuous dasatinib (60 mg/m2 orally twice daily) and everolimus (3 mg/m2 orally once daily) at four academic institutions. We treated four patients with recurrent grade II-IV glioma and two patients with high-grade glioma after initial radiation (Table 1). All patients tolerated the combination with the exception of rash (n=4/6), mucositis (n=1) and fatigue (n=1); none of which required dose reduction. In recurrent patients (n=4), median progression-free survival (PFS) was 3 months (range 2-5 months) and median overall survival (OS) was 8.5 months (range 3-16 months and ongoing) (Figure 4A-B); although some patients had additional concurrent therapies (Table 1).

**Table 1.** Human glioma patient characteristics. Patient characteristics are shown for patients that underwent treatment with dasatinib and everolimus across four academic institutions (n=6).

| Recurrent | Patient ID | Age | Diagnosis | Molecular | Cycles on therapy | PFS on therapy | Best Response | Overall survival from Recurrence | Toxicity/Other notes |
| --- | --- | --- | --- | --- | --- | --- | --- | --- | --- |
|  | UMPED44 | 7 | Recurrent diffuse midline glioma, H3 wildtype (thalamus) | *PDGFRA* amplification, *TP53* (R65*), *CDKN2A* loss | 3 cycles | 3 months | SD (cycle 2) | 8 months | Skin rash (did not require dose reduction) |
|  | UMPED52 | 4 | Recurrent diffuse intrinsic pontine glioma (DIPG), H3 wildtype | *PDGFRA* mutation (D842V), *ACVR2B* (E347*) | 4 cycles | 5 months | SD (cycle 2+4) | 9 months | No toxicity noted |
|  | UMPED68 | 19 | Recurrent diffuse astrocytoma, H3 wildtype (frontal-temporal) | *PDGFRA* indel (S1045fs), *EGFR* (A289D, P596L), *PTPN11*(R498W) | 5 cycles | 3 months | SD (cycle 2) | 16 months (ongoing) | Skin rash (did not require dose reduction); received focal radiation between cycles 3 and 4 |
|  | SCH01 | 10 | Recurrent high-grade glioma, H3 wildtype, secondary to ependymoma radiation (posterior fossa) | *PDGFRA* amplification, *KIT* amplification, *CDK2NA* deletion, *MTAP* deletion, *GAB2* amplification | 2 cycles | 2 months (presumed , no further imaging) | Not assessed (no further imaging) | 3 months | Mild rash and fatigue (did not require dose reduction); oral etoposide added after first cycle |
| Adjuvant (newly diagnosed) | Patient ID | Age | Diagnosis | Molecular | Cycles on therapy | PFS on therapy | Best Response | Overall survival from Recurrence | Toxicity/Other notes |
|  | NYU01 | 15 | Metastatic high-grade glioma, H3 wildtype, secondary to leukemia (multiple hemispheres) | *PDGFRA* mutation (D842V) and amplification, *KIT* mutation (D579N) and amplification | 5 cycles (ongoing) | N/A (not progressed) | SD/minor response (cycle 3) | 8 months (ongoing) | No toxicity noted; therapy started adjuvantly after initial radiation |
|  | UCSF01 | 8 | Diffuse intrinsic pontine glioma (DIPG), H3K27M-mutant | *PDGFRA* up-regulation (RNA); *H3F3A* (K27M); *ATRX* (R2111L); *MTOR* (E1799K); *PI3K* (E545K); *PPM1D* (C487*) | 3 cycles | 3 months | SD (cycle 2) | 11.8 months | Mucositis, oral pain, rash (did not require dose reduction); oral metformin and etoposide treatment given concurrently |

Additionally, to measure CSF and plasma levels of dasatinib with and without concurrent everolimus, we collected paired CSF/plasma samples from two patients after one week of dasatinib mono-therapy and then after two cycles of dual therapy with dasatinib and everolimus, according to institutional IRB-approved protocol (Supplemental Figure 10A). Patient UMPED44 demonstrated an increase in both plasma (11.2 ng/ml to 31.5 ng/ml) and CSF (0.44 ng/ml to 0.91 mg/ml) dasatinib levels (Figure 4C and Supplemental Figure 10A-C). UMPED52 demonstrated an increase in plasma (26.4 ng/ml to 165 ng/ml) dasatinib levels, but CSF samples were unable to be collected (Supplemental Figure 11A-D).

### P-gp expression in human HGG RNA-seq datasets

As P-gp is expressed both in brain and tumor cells, and overall expression at protein level was reduced in our mouse HGG models after sustained treatment with everolimus, we were interested to explore if P-gp expression correlated with clinical or genomic features in human datasets. Previous data has shown correlation between P-gp RNA and protein expression in human liver samples, with both predicting organ penetration of a Pgp-effluxed compound (29). We analyzed RNA-seq data from pediatric HGGs within PedcBioPortal, [Pediatric HGG ICR London dataset, n = 247 with RNAseq data (2)] and found that P-gp expression was highest in: (a) pediatric hemispheric HGG, (b) tumors with wild type *H3F3A* (gain of function mutations are found in half of pediatric HGG), and (c) young adult age patients (age 10-15) (Figure 5A). Importantly, P-gp expression was higher in pediatric HGG with *PDGFRA* alterations (Figure 5A, n= 147). In analysis of adult HGGs (cBioPortal, TCGA dataset, n = 585) (30), no clinical parameters showed correlation with P-gp expression, including *PDGFRA* status, MGMT status and age at diagnosis (Supplemental Figure 12A). Future studies involving P-gp IHC of human tissue are needed to further explore possible correlations between P-gp expression and clinical or genomic features.

## DISCUSSION

Our study illustrates that everolimus improves the efficacy of dasatinib in the treatment of PDGFRα-driven glioma. While response attribution is difficult for our heterogeneous consecutive cases, our early clinical experience with this combination demonstrated excellent tolerance of the regimen and overall survival from progression that compares favorably to historical controls (27, 28). To our knowledge, the use of everolimus co-treatment with dasatinib had not previously been tested in glioma tumor models or human patients. More broadly, our results demonstrate that TKI treatment targeted to alterations in HGG may have renewed clinical relevance when combined with mTOR inhibitors.

Importantly, our murine IUE tumor model experiments demonstrate that everolimus and dasatinib display improved combinatorial activity in the treatment of PDGFR-driven glioma cells. This may be due to the presence of dual inhibition within the MAPK/mTOR cell proliferation pathway: with everolimus reducing residual mTOR activation from dasatinib treatment alone (5, 31) (Figure 5B). Attribution to tumor cell toxicity from improved intra-cellular dasatinib concentration vs. direct mTOR inhibition is difficult to fully distinguish. However, we found no in vitro P-gp inhibition with everolimus treatment at low dose (10 nM), while our treatment studies show significant contribution of everolimus to reduced tumor cell viability at doses as low as 1 nM. Murine PPK cells did display dasatinib/ everolimus synergism via the Chou-Talalay median-effect method for drug combination analysis (20) (Figure 2E). Future mechanistic studies may further clarify whether the benefit is additive (vs synergistic).

In our in vivo model, the administration of everolimus increased tumor concentrations of dasatinib above therapeutic (IC50) levels, and this increase required sustained (>24 hours) everolimus exposure. This requirement may be related to the time required for protein-level changes (i.e. reduced P-gp expression and/or activity) to occur. Interestingly, dasatinib concentrations were raised more prominently in the CNS tumor samples than the brain itself. This may be due to the disruption of the BBB in high-grade tumors, allowing for greater dasatinib influx (32). Indeed, our murine tumor demonstrated high-grade features that may influence BBB integrity, including necrosis and vascular proliferation (Supplemental Figure 1A).

A concern with P-gp inhibition strategies that remains in our study is whether the inhibitor itself can get into the CNS at high enough concentrations to achieve P-gp inhibition (33). Our data showed a concentration of everolimus required for P-gp inhibition that was achievable in the plasma (∼10 uM) and brain (∼1 uM) of our mouse models, but may be more difficult to achieve in human brains with current clinical trough goals (5-15 ng/mL, or

∼5-15 nM). Interestingly, early adult phase 1 studies of everolimus studied weekly higher dosing of everolimus (up to 70 mg), which resulted in mean peak levels of everolimus of 175 ng/ml (190 nM), and tolerability similar or better than daily 10 mg dosing (34). Future studies may clarify whether everolimus inhibits P-gp at this dosing or whether higher dosing of everolimus should be re-considered. P-gp also promotes efflux on the luminal side of blood vessels (Figure 5B), and so future clinical efforts to achieve Pg-P inhibition may be based on plasma (rather than brain parenchyma) concentrations of everolimus. At a minimum, based on our in vitro models, everolimus should contribute to synergistic cytotoxicity of PDGFR-driven HGG in clinically relevant (∼low nM) concentrations of everolimus and dasatinib.

While everolimus may help prevent efflux of dasatinib out of brain parenchyma and tumor cells, the efficacy of dasatinib will ultimately rely on the cells’ sensitivity to it, which may be variable based on the type of *PDGFRA* alteration expressed by the cells. The *PDGFRA* alteration in each glioma can strongly contribute to the sensitivity to dasatinib (7). Our PPK cells carry the recurrent *PDGFRA* D842V mutation (as did patient UMPED52) and displayed nM reduction in proliferation to dasatinib. Previous studies have shown mixed sensitivity of the D842V mutant to dasatinib, including reduced sensitivity to dasatinib in *PDGFRA* D842V-mutant normal human astrocytes (NHAs) (7). However, nanomolar reduction in D842V-mutant kinase activity has been demonstrated in gastro-intestinal stromal tumor (GIST) tumors (35). Of note, our IUE-based PPK cells retain endogenous, wild-type *PDGFRA*, which may contribute to tumor proliferation and dasatinib sensitivity. Future studies will continue to clarify the responsiveness of various *PDGFRA* alterations to dasatinib sensitivity, but our data demonstrates promise of this dual therapy in both *PDGFRA*-mutant and amplified gliomas.

Our clinical case series (n=6) confirms feasibility for this dual therapy in this very high-risk patient population. Patients tolerated this combination without significant adverse events. Efficacy is difficult to fully assess due to heterogeneity and concurrent therapies in this group, but the median OS (8.5 months) in the four recurrent pediatric glioma patients was slightly higher than historical controls for relapsed pediatric glioma (four to six months) (27, 28). While we did find an increase in CSF dasatinib levels after everolimus co-administration in UMPED44, the overall CSF dasatinib concentration (∼2 nM) still remained near or below the IC50 of most human PDGFRα-driven glioma cell cultures. We are using CSF as the best proxy for dasatinib concentrations within the tumor, and it may not be an accurate reflection of tumor concentration. Indeed, in our murine model, cortex concentrations of dasatinib were consistently lower than tumor concentrations. Future patient data will help clarify the CSF retention of dasatinib, and future clinical studies may consider further dose escalation of dasatinib.

Thus far, targeted treatments for PDGFRα-driven HGG have failed to improve outcomes in both adult and pediatric patients. This shortcoming has exposed the need for new precision medicine approaches for these patient population. Our study confirms that dasatinib effectively inhibits *PDGFRA*, and the sustained co-administration of dasatinib and everolimus allows for additional combinatorial efficacy and the potential for improved CNS penetration. The combination of dasatinib and everolimus represents a new avenue for adult and pediatric HGG treatment, and the co-administration of these drugs re-opens the possibility of dasatinib being an effective therapy for PDGFRα-driven glioma.

## METHODS

### Murine model of high-grade glioma using intra-uterine electroporation

In utero electroporation (IUE) was performed using sterile technique on isoflurane/oxygen-anesthetized pregnant CD1 females at embryonic stage E 13.5 (cortex), using established methodology (36). All tumors for this study were generated by injecting plasmids either in the lateral ventricle (forebrain) or in the fourth ventricle (hindbrain). In this study we injected the following four plasmids together: [1] PBase, [2] PB-CAG-DNp53-Ires-Luciferase (dominant negative TP53 or TP53 hereafter), [3] PB-CAG-*Pdgfra*D824V-Ires-eGFP (“*PDGFRA* D842V”), and [4] PB-CAG-H3.3 K27M-Ires-eGFP (“H3K27M”), referred to as “PPK” model.

Following anesthesia induction, Carprofen was administered subcutaneously for additional analgesia. Uterine horns were exposed through a 1 cm incision, and individual embryos were digitally manipulated into the correct orientation for intraventricular injection. A pulled capillary needle was loaded with endotoxin-free DNA and Fast Green dye (0.05%, Sigma-Aldrich) for visualization, and a microinjector was used to inject either the lateral or fourth ventricles with the DNA-dye mixture. 3-4 plasmids were injected simultaneously, each at a final concentration of 1 μg/μl and 1-2 μl of total solution was injected per embryo. DNA was then electroporated into cortical neural progenitors using 5 mm tweezertrodes (BTX), applying 5 square pulses at 35 V, 50 ms each with 950 ms intervals. The embryos were then returned into the abdominal cavity, the muscle and skin were sutured and the animal was monitored until fully recovered from the procedure.

After delivery, the efficacy of plasmids uptake was monitored by bioluminescence with the IVIS Spectrum (tumors express luciferase). Pups that did not display bioluminescence were euthanized, which occurred in about 5 to 10 percent of the pups.

After three weeks, juveniles were weaned and separated by gender. The mice with positive signal were monitored for tumors every day by observation and biweekly bioluminescence imaging on an IVIS Spectrum imaging system until the signs of intracranial tumor burden ensued.

### Generation of IUE primary HGG cell cultures

Mouse HGG primary cell cultures were generated by harvesting IUE tumors at the time of euthanasia. Tumors were located by green fluorescent protein (GFP) expression under an epi-fluorescent microscope (Olympus CKX41) at the time of resection. The tumor mass was gently homogenized and dissociated with non-enzymatic cell dissociation buffer (Gibco). Cell suspension was filtered through a 70 μm cell strainer, centrifuged at 300 × g for 4 min, and re-suspended into 7 ml of Neurobasal-A Medium (1x) Base media (Invitrogen, 500 mL). Each base stock of media was supplemented with 10 mL B27 with vitamin A, 5 mL N2, 5mL Sodium Pyruvate (1 mM), 5 mL non-essential amino acids (100 uM), 7.5 mL L-glutamine (2 mM), 1mL Normocin, 5 mL Antibiotic-Antimycotic, and 1μL EGF (20ng/mL) per 1 mL media every three days.

### In vitro *treatment and synergism calculations*

IUE-generated PDGFRα-driven HGG primary cell culture (PPK) were cultured in conditions as described above. Cell viability in vitro was monitored either by XTT Cell Proliferation Assay kit (Cayman Chemical) utilizing the included protocol, or bioluminescence readings collected using the Synergy HTX Multi-Mode micro plate-reader (BioTek). Additionally, cell proliferation was monitored via IncuCyte Zoom live-cell analysis. Through use of the IncuCyte Zoom live imager, the growth of 3,000 PPK primary cells plated in 96 wells was monitored by units of average green fluorescent intensity (GCU × um2). PPK primary cell culture was grown with 200 μL of media and 1 μL of PDGF-AA and PDGF-BB at working concentration of 10 ng/mL in each 96 well (Shenandoah Biotechnology, Inc.). All in vitro treatments were completed with research-grade dasatinib and everolimus (Selleck). UMPED58 is a primary human cell line generated from a 7-year-old male with DIPG at autopsy. At diagnosis, tumor was sequenced and found to carry *H3F3A K27M, ATRX pQ119\**, and *PDGFRA, KIT* and *KDR* amplifications. UMPED05 is a primary cell line generated from a 2-year-old female with thalamic HGG at autopsy with *PDGFRA* amplification, as previously described by Koschmann et al. (2016) (5). UMPED58, UMPED05 and KNS42 were grown in 10 mL L-glutamine (200mM), 5 mL 100x penicillin streptomycin (pen strep), 10% fetal bovine serum. KNS42 was kindly provided by Dr. Alan Meeker (Johns Hopkins University, Baltimore, MD). Mayo 154C, Mayo 64VF, Mayo 200VF were derived from PDX models kindly provided by Dr. Jann Sarkaria (Mayo Clinic College of Medicine, Rochester, MN) (37), and DIPG-XIII-P, kindly provided by Dr. Michele Monje (Stanford University, Stanford, CA) (38). All synergism calculations were performed by the Chou-Talalay method for drug combination based on the median-effect equation (20).

Briefly, we entered normalized proliferation assay data for combinations of no treatment and four serial dilutions of each compound (D1 and D2) to generate linear regression curves for each compound, resulting in generation of theoretic doses (Dx1 and Dx2). Comparison of actual dose (D1) and theoretic dose (D1/Dx1)+(D2/Dx2) provides a combination index (CI). The Chou-Talalay theorem offers quantitative definition for additive effect (CI = 1), synergism (CI < 1), and antagonism (CI > 1) in drug combinations (20).

### Murine IUE HGG treatment studies

Mice harboring IUE-generated PPK HGG tumors were treated with everolimus and/or dasatinib. Mice were treated when tumors reached logarithmic growth phase (minimum 2 × 10^5^ photons/sec via bioluminescent imaging), and confirmation was made that treatment groups had equivalent average luminescence at time of treatment.

Mice litters from each experimental group were randomized to treatment with: (A) 10 mg/kg dasatinib or (B) 5 mg/kg everolimus alone (10% dasatinib or everolimus suspended in DMSO, 80% ultra-pure water, 10% Tween-80), (C) a combination group which received 10 mg/kg dasatinib and 5 mg/kg everolimus, or (D) control treatment (10% DMSO, 80% ultra-pure water, 10% Tween-80). Initial studies with the high-dose combination (25 mg/kg dasatinib and 10 mg/kg everolimus) were associated with early treatment-related morbidity/mortality; therefore, this arm was excluded from further studies. Mice were treated every other week on this schedule until morbidity.

Animals displaying symptoms of morbidity after treatment were euthanized for immunohistochemical or pharmacodynamic/ pharmacokinetic (PD/PK) tumor. Some mice (n=3) were treated for an initial treatment dose of their assigned arm 4 hours prior to euthanization. For intra-tumor concentration studies, all doses of everolimus were administered to mice 24 hours prior to dasatinib administration, consistent with previous methods used for in vitro analysis of mTOR/MAPK regulation after treatment with everolimus (10). For immunohistochemistry analysis, mice were perfused with Tyrode’s Solution followed by 4% paraformaldehyde fixative solution to preserve the structures of the brain.

### Pharmacokinetic analysis

#### Mouse PK sample procurement

Drug administration to non-tumor bearing C57/Bl6 mice for PK studies was performed by oral gavage administration of everolimus two hours prior to a tail vein injection of dasatinib. At one, two, four and seven hours after the dasatinib injection, the mice were isoflurane/oxygen-anesthetized and 500 uL to 1 mL of blood was drawn from the apex of the heart within the mouse’s enclosed cavity. Immediately, the withdrawn blood was centrifuged within a microvette EDTA coated conical tube for 10 minutes at 10,000 RPM, and the plasma was separated and stored at -80°C until PK analysis was performed.

Following the blood draw, the mouse was sacrificed and the brain, brain stem, and/or tumor were extracted separately and stored at -80°C until PK analysis was performed.

#### Chemicals and reagents

For PK studies, dasatinib powder was procured from Sigma-Aldrich. Liquid chromatography–mass spectrometry (LC-MS) grade acetonitrile was purchased from Sigma-Aldrich. Formic acid (98%; LC-MS grade) was obtained from Fluka. A Milli-Q water system from Millipore was used to obtain ultrapure deionized water.

#### Stock solutions, working solutions, and quality control samples

Dasatinib and the internal standard were individually weighed and dissolved in acetonitrile to stock solutions and then stored at −20°C. The dasatinib stock solution was then diluted with acetonitrile to a series of working solutions from 2.5 to 5,000 ng/mL The quality control working solutions at low, medium, and high concentrations were prepared using a separately prepared stock solution. For sample preparation, the dasatinib stock solution was diluted to 1,000 ng/mL with acetonitrile. Quality control samples were evenly distributed among samples from each batch.

#### Sample preparation

Plasma (40 µL) was dispensed into a Fisher Scientific 96-well plate, to which 40 µL of ice-cold acetonitrile (100%) and 120 µL of internal standard solution (1000 ng/mL) were added. Next, the plate was vortexed for 10 minutes. The plate was then centrifuged at 3500 revolutions per minute (RPM) for 10 minutes at 4°C to precipitate the protein. LC–tandem mass spectrometry (LC-MS/MS) was used to analyze 5 µL of the supernatant. The plasma samples were sonicated prior to being transferred to the 96-well plates. Tissue samples were weighed and suspended in 20% acetonitrile (80% water; 1:5 wt/vol). The samples were then homogenized four times for 20 seconds each time at 6,500 RPM in a Precellys Evolution system. For LC-MS/MS analysis, the dasatinib in brain tissue homogenates were extracted from the samples in the same manner as the dasatinib in plasma. Prior to extraction, samples that were above the upper limit of qualification were diluted with the same matrix. Calibrator-standard samples and quality control samples were prepared by mixing 40 µL of blank bio matrix, 40 µL of working solution, and 120 µL of internal standard solution.

#### Calibration curve

Analytical curves were made with 12 nonzero standards by plotting the peak area ratio of dasatinib to the internal standard vs the concentration. The curve was created with linear regression and weighted (1/X2). The correlation coefficient demonstrated the linearity of the relationship between peak area ratio and concentration.

#### Liquid chromatography tandem–mass spectrometry

The concentrations of dasatinib were determined with a Sciex AB-5500 Qtrap mass spectrometer with electrospray ionization source, interfaced with a Shimadzu high-performance LC system. The LC-MS/MS system was controlled with Analyst Software version 1.6 from Applied Biosystems; this was also used for acquisition and processing of data. Separation was performed on a Waters Xbridge C18 column (50 × 2.1 mm ID, 3.5 µm); the flow rate was 0.4 mL/min. A (100% H2O with 0.1% formic acid) and B (100% acetonitrile with 0.1% formic acid) comprised the mobile phase. The gradient began with 5% B for 30 seconds and then linearly increased to 99% B at 2 minute and then reduced to 5% B at 4.1 minutes to 5.5 minutes with a runtime of 6 minutes in total. The mass spectrometer was operated in positive mode; multiple reaction monitoring was used for analysis. The Q1 m/z and Q3 m/z was 487.9 and 401.1, respectively.

### Human treatment studies

All patients received dasatinib 60 mg/m2 orally twice daily and everolimus 3 mg/m2 orally once daily continuously, with titration of dosing to maintain an everolimus trough level of 5-15 ng/mL. Four patients were treated off-therapy (UMPED68, SCH01, NYU01, and UCSF01) and two patients (UMPED44 and UMPED52) were treated on an IRB-approved clinical trial. Enrollment required CLIA-certified confirmation of a DNA (mutation, amplification) or RNA (fusion) alteration in a PDGF-related gene (*PDGFRA, PDGFA, PDGFRB, PDGFB*). Paired CSF/plasma samples (before and after addition of everolimus) were collected from enrolled patients when clinically feasible. Cycles were repeated every 28 days for up to 24 cycles. CSF and serum dasatinib pharmacokinetics were performed by the University of Michigan Pharmacokinetic Core Facility (see supplemental methods) to determine whether blood-CSF permeability of dasatinib is impacted by dual therapy with everolimus. Prior to the first cycle, dasatinib was taken as mono-therapy for one week followed by CSF collection in order to establish a mono-therapy dasatinib CSF level. After the second cycle, CSF was collected to establish a dual-therapy dasatinib level (while on everolimus). Additionally, all CSF samples were assessed for cell-free tumor DNA (cf-tDNA) using previous methods (39), when sample remained.

### Human HGG clinical sequencing and PDGFRA alteration confirmation

For confirmation of *PDGFRA* status in human HGG patients, tumor (FFPE or frozen) and normal (cheek swab) samples were submitted for whole exome (paired tumor and germline DNA) and transcriptome (tumor RNA) sequencing. Clinically-integrated sequencing was performed after enrollment in the University of Michigan IRB-Approved Pediatric MiOncoseq Study using standard protocols in adherence to the Clinical Laboratory Improvement Amendments (CLIA). All somatic mutation information generated from this study has been uploaded to dbGaP (https://www.ncbi.nlm.nih.gov/gap) under accession number phs000673.v2.p1.

### Human P-gp expression

Pediatric and adult human P-gp expression dataset analysis was performed using RNA-seq data accessed from PedcBioPortal. For pediatric HGG, data was accessed from the Pediatric HGG ICR London (2). We performed comparisons of P-gp expression by location of tumor, H3 (*H3F3A* or *HIST1H3B*) mutation status, *PDGFRA* mutation status, and age. For adult HGG, data was accessed from The Cancer Genome Atlas dataset (30) (30). For TCGA (adult HGG) P-gp RNA, we compared P-gp expression levels by *PDGFRA* mutation status, age and methylguanine-DNA methyltransferase (MGMT) status.

### Statistics

For comparison of dose-response curves using DNA-damaging agents, a nonlinear regression curve [log (agent) vs. normalized response with variable slope] was generated. Statistical significance in all experiments was defined as a two-sided *P* ≤ 0.05. All analyses were conducted with GraphPad Prism software (version 7.00). Expression level of p-Src and p-S6 were quantified using IHC performed on mouse brain sections treated with everolimus and/or dasatinib. Four randomly selected JPEG images were captured from each IHC slide at 12.3x magnification on the Aperio image scope and quantified using ImageJ software. ImageJ software was also used to quantify IHC and western expression level as well as PPK neurosphere area in squared pixel unit. Comparisons between treatment groups in experiments with only two groups were completed using a two-tailed Welch’s t-test. Comparisons between treatment groups in experiments with more than two groups were completed using a Tukey’s multiple comparison test or a Dunnett’s multiple comparisons test. Survival curve comparisons between treatment groups were performed using the log-rank test.

### Study Approval

#### Animal studies

All animal studies were conducted according to the guidelines approved by the University Committee on Use and Care of Animals (UCUCA) at the University of Michigan.

#### Human studies

Four patients were treated off-therapy (UMPED68, SCH01, NYU01, and UCSF01) after institutional IRB-approved protocols for molecular tumor assessment. Two patients (UMPED44 and UMPED52) were treated on an IRB-approved clinical trial with dasatinib and everolimus for children and adults with newly diagnosed (post-radiation) high-grade (grade III-IV) glioma or recurrent grade II-IV glioma with *PDGF* alterations (NCT03352427).

## AUTHOR CONTRIBUTIONS

ZM, VNY, BM, RTC, CCT, and CKo performed in vitro experimental design, experiments and data analysis. ZM, VNY, BM, TG, TNP, TY, RTC, CCT, JRC, DRW, and JNS performed in vivo experimental design, experiments and data analysis. ZM, VNY, RS, BLM, KW, AKB, SS, HJLG, AP, IW, ML, PLR, SESL, SV, TN, CG, CKl, SM, CK-S, AMC, RM, and CKo performed clinical/human studies. ZM, VNY, MPP, BLM, and CKo completed pharmacokinetic analysis. ZM, VNY, BLM, JRC, CKo, RS, RTC, and SV completed manuscript preparation. All authors discussed and reviewed the manuscript and approved the manuscript for publication.

## Data Availability

De-identified human sequencing data will be made publically avilable according to established practices of University of Michigan MCTP/MiOncoseq practice.

## ACKNOWLEDGMENTS

The authors thank the patients and their families for participation in this study.

